# Multi-Omic Graph Diagnosis (MOGDx) : A data integration tool to perform classification tasks for heterogeneous diseases

**DOI:** 10.1101/2023.07.09.23292410

**Authors:** Barry Ryan, Riccardo E. Marioni, T. Ian Simpson

## Abstract

Heterogeneity in human diseases presents challenges in diagnosis and treatments due to the broad range of manifestations and symptoms. With the rapid development of labelled multi-omic data, integrative machine learning methods have achieved breakthroughs in treatments by redefining these diseases at a more granular level. These approaches often have limitations in scalability, oversimplification, and handling of missing data. In this study, we introduce Multi-Omic Graph Diagnosis (MOGDx), a flexible command line tool for the integration of multi-omic data to perform classification tasks for heterogeneous diseases. MOGDx is a network integrative method that combines patient similarity networks with a reduced vector representation of genomic data. The reduced vector is derived from the shared latent embedding of a multi-modal encoder and the combined network is fed into a graph convolutional network for classification. The multi-modal encoder and graph convolutional network are trained simultaneously making a fully supervised pipeline. MOGDx was evaluated on three datasets from the cancer genome atlas for breast invasive carcinoma, kidney cancer, and low grade glioma. MOGDx demonstrated state-of-the-art performance and an ability to identify relevant multi-omic markers in each task. It did so while integrating more genomic measures with greater patient coverage compared to other network integrative methods. MOGDx is available to download from https://github.com/biomedicalinformaticsgroup/MOGDx. Overall, MOGDx is a promising tool for integrating multi-omic data, classifying heterogeneous diseases, and interpreting genomic markers.

## Introduction

Heterogeneity in human diseases is a pertinent yet difficult issue that can confound the analysis of clinical trials, genetic association testing, drug responses, and intervention strategies. Heterogeneous diseases encompass any single disease with a broad range of manifestations or symptoms. Redefining such diseases through sub-type classification, symptomatic grading or similar has the potential to uncover new treatments, re-purpose old treatments or identify intervention strategies. This approach has already been shown to improve patient outcomes in a number of diseases (Brodlie et al., 2015; Sosman et al., 2012). Performing classification tasks with heterogeneous diseases is a complex problem often requiring analysis of multiple types of data of varying scale and complexity, as such it needs analytic frameworks that are flexible and scalable. The use of Artificial Intelligence (AI) has emerged as a popular method to solve this problem and has been facilitated by the development of high-throughput sequencing technologies. Such technologies have made various types of biological data, coined ‘omic’ data, available. This increased availability of omics has led to many novel bioinformatic analytical tools for individual omics. This individual omic focus has yielded positive outcomes, however information from different omics are rarely compared. Identifying which is the most informative omic measure, and if the information captured in different omics overlap or are orthogonal, is largely unknown. We hypothesise that while an individual omic can provide a single biological measure, the integration of multiple informative omics could capture multiple biological measures and increase classification accuracy in heterogenous diseases. Therefore, an analytical tool which can integrate multiple omic measures and perform heterogeneous disease classifications could significantly improve patient outcomes in this area.

There is an increasing number of methods which integrate multi-omic data in both the supervised and unsupervised classification space. There exists two main taxonomies for data integration, which can be broadly categorised as input data-fusion and output data-fusion, although no gold standard method exists. Input data-fusion methods combine data sources into a single dataset prior to analysis, while output data-fusion methods analyse each dataset separately and combine the results. Input data-fusion methods estimate an embedding which projects datasets into a shared latent space which minimizes variance between datasets while maximising individual variability within each data set (Gliozzo et al., 2022). For example, Lock et al. (2013) achieved state-of-the-art performance on characterization of tumour types from the Breast Invasive Carcinoma (BRCA) dataset in The Cancer Genome Atlas (TCGA)(https://www.cancer.gov/tcga). Their analysis was able to effectively uncover both individual and joint data structures, resulting in better interpretation while also improving unsupervised classification results on the BRCA dataset. In its simplest form, output data-fusion resembles ensemble methods, whereby an independent analysis of each dataset is performed, and the results combined using an aggregation technique. An example of this is presented by Phan et al. (2016) who use a stacked-generalisation model on an ovarian dataset in TCGA.

These methods show that classification performance is increased when multiple modalities are considered, however they often scale poorly, are overly simplistic or do not take into account the cross correlation between modalities (Gliozzo et al., 2022; Wang et al., 2021). The use of a network taxonomy for multi-omic data integration has risen in popularity recently. The advantage of networks is that they are easily integrated and can readily handle missing data. Gliozzo et al. (2020) and Li et al. (2015) show that representing data as a Patient Similarity Network (PSN) can retain information and have superior or competitive performance compared to standard Euclidean methods for a single modality. netDx, developed by Pai et al. (2019), uses ridge regression and label propagation algorithms to integrate and perform ranked classifications on PSN’s. Wang et al. (2021) define each modality as a single PSN, perform classification using a Graph Convolutional Network (GCN) and concatenate predictions into a cross-omic correlation tensor before making final label predictions. Li et al. (2022) perform classifications using a GCN on integrated PSN’s. These methods are novel strategies for the integration of network data at the input and output space, however, they don’t leverage the full advantages of representing data as a network. Current network methods cannot handle patients missing one or more omic measures, and methods can only handle a fixed number of modalities.

Hence, we introduce Multi-Omic Graph Diagnosis (MOGDx), a flexible tool for the integration of multi-omic data to perform classification tasks for heterogeneous diseases. The MOGDx pipeline integrates omic data into a single PSN before performing classification using an Graph Neural Network (GNN) algorithm. Each omic measure undergoes pre-processing steps to extract the most informative features. These features are used to inform the PSN for each individual omic measure. Similarity Network Fusion (SNF)(Wang et al., 2014) is performed to integrate the PSN’s into a single network. This network, along with the omic measures, are passed into the Graph Convolutional Network with Mulit-Modal Encoder (GCN-MME) model. Each unprocessed omic measure is passed through the Mulit-Modal Encoder (MME) for supervised dimensionality reduction. The MME consists of a two layer encoder which compresses each modality into a reduced latent space. This latent space is then decoded into a fixed shared vector, such that each modality is encouraged to learn the same reduced representation. Each vector has a corresponding patient node in the fused PSN and the two are combined into a single GNN model, namely GCN in this instance. Backpropagation is performed through both the GCN and MME for a fully supervised training pipeline. The performance of MOGDx is benchmarked on the BRCA, Low-Grade Glioma (LGG) and Pan Kidney Cohort (KIPAN) datasets, with state-of-the-art performance demonstrated. MOGDx is the first tool of its kind which can handle missing patient data as well as any number of data modalities or omic measures. We show the benefit of integrating multiple modalities and the importance of representing data as a PSN. We demonstrate that MOGDx can identify informative omic measures as well as important omic markers relating to the targeted biomedical problem.

## Results

### Pipeline of MOGDx

We present MOGDx, a pipeline for the supervised classifications of patients with heterogenous diseases (Figure 1). MOGDx takes as input any number of omic measures such as genomic, transcriptomic and proteomic datasets. The raw data is processed into omic measure matrices, with each row corresponding to a patient and each column a feature of that measure. First, depending on the omic dataset (see Methods), either differential expression or penalised logistic regression is performed to identify important features of that dataset. These features are used to inform a weighted similarity matrix calculated using Pearson correlation. A PSN is then constructed using the K-Nearest Neighbours (KNN) algorithm. SNF is performed to fuse the PSN’s into a single network.

**Fig. 1:**
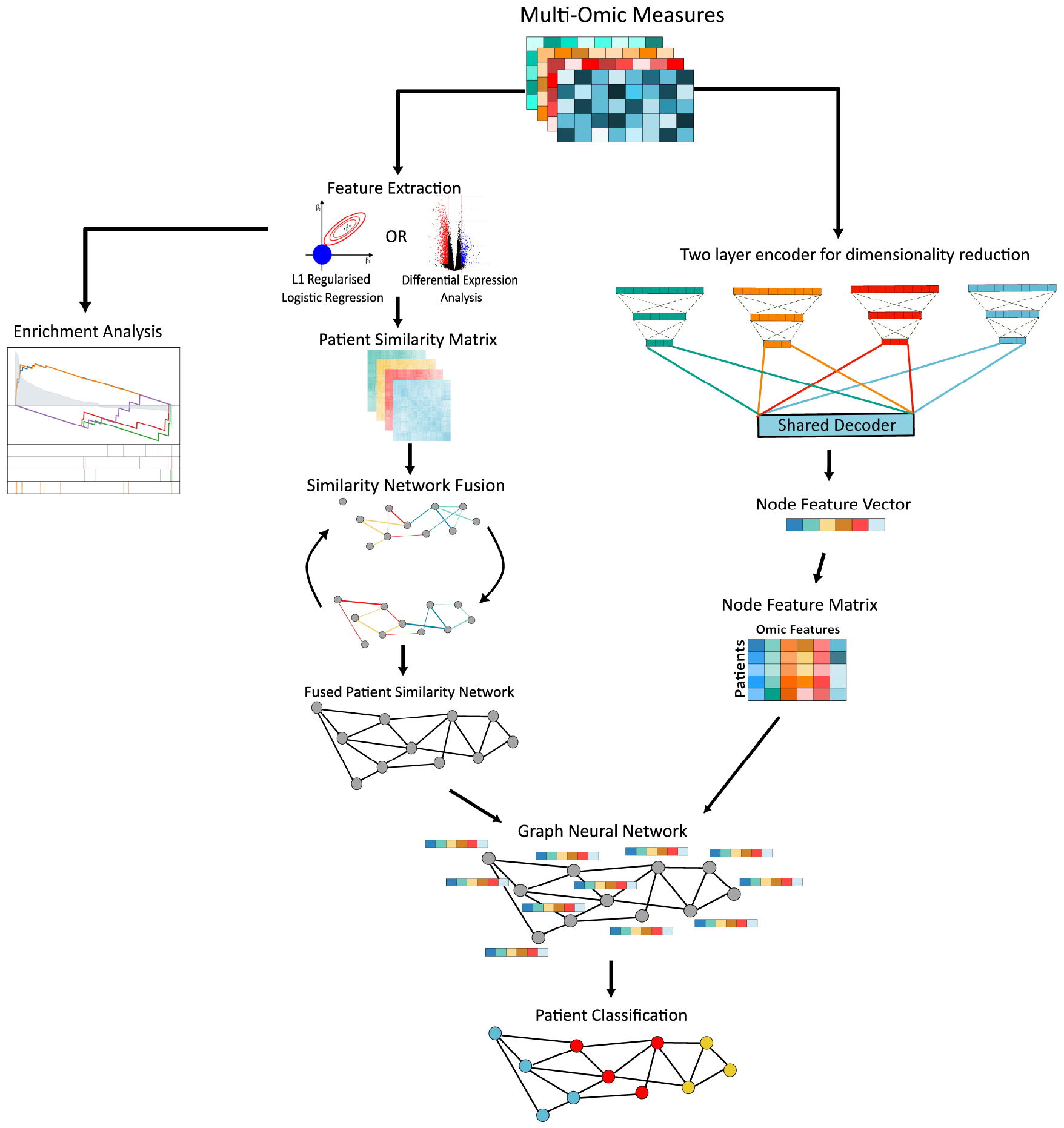
Pipeline of MOGDx. MOGDx takes any number of omic measures as input. Feature extraction is performed to maximise similarities between patients. Each patient similarity matrix is converted to a network and these patient similarity networks are fused using SNF. The fused PSN and the unprocessed omic measures are input into the GCN-MME model for training. The MME provides a shared reduced node feature vector for each patient node in the network. Training is performed by back propagating through the entire GCN-MME model for a fully supervised pipeline

From here, the fused PSN and the omic datasets are input into the GCN-MME, the architecture of which is shown in Figure S1. Each omic measure is encoded using a two layer encoder. The first layer of the encoder is of fixed length, with the second layer being tuned to each modality by performing a hyperparameter search. The compressed encoded layer of each modality is then decoded to a shared latent space using mean pooling. The goal of this step is to encourage each modality to learn the same latent space. The shared latent space is the node feature matrix, required for training the GCN, with each row forming a node feature vector. This node feature vector corresponds to a single node in the PSN. The node feature matrix and fused PSN are combined and input into a GNN. The dimension of the shared latent space was tuned through hyperparameter searching. The optimal hyperparameter values for the best performing modalities in each dataset are given in Table S1.

A GCN architecture was implemented using the Deep Graph Library (version 1.1) in Python with a PyTorch backend. The GCN implemented consisted of two layers with intermediate relu activation and batch normalisation. The input dimension to the GCN is the same dimension as the node feature vector. The output dimension of the second layer aligns with the number of classes for that classification task. The weights of both the GCN and MME are learned jointly by performing backpropagation through the entire GCN-MME. This methodology integrates the predictive power of PSN’s with a reduced representation of classical omic characteristics. MOGDx obtains state-of-the-art predictive performance on the integration of network and vector characteristics, exhibiting the benefit of including both. MOGDx is a command line tool for the supervised classification of heterogeneous diseases, which can be used for a wide range of biomedical applications.

### Datasets

The performance of MOGDx is benchmarked on three different TCGA datasets, BRCA PAM50 sub-type classification, grade classification in LGG and KIPAN for kidney type classification. Data was downloaded using the TCGABiolinks(Colaprico et al., 2023) Bioconductor package (version 2.28.3). All modalities available in the TCGA database were included, resulting in 5 types of omics data used for classification. The omic data types available are mRNA expression (mRNA) data, micro RNA expression (miRNA) data, DNA methylation (DNAm) data, Reverse Phase Protein Array (RPPA) data and Copy Number Variation (CNV) data. All patient samples were utilised irrespective if they were available in only one or all datasets, with specific details reported in Table 1. Raw feature count is the total number of features available per modality. Processing was performed to remove features which were mostly missing or had a standard deviation of 0. The processed features were directly inputted into the GCN-MME. Further processing was performed to extract the most predictive features, which were used to construct the PSN’s.

**Table 1.**
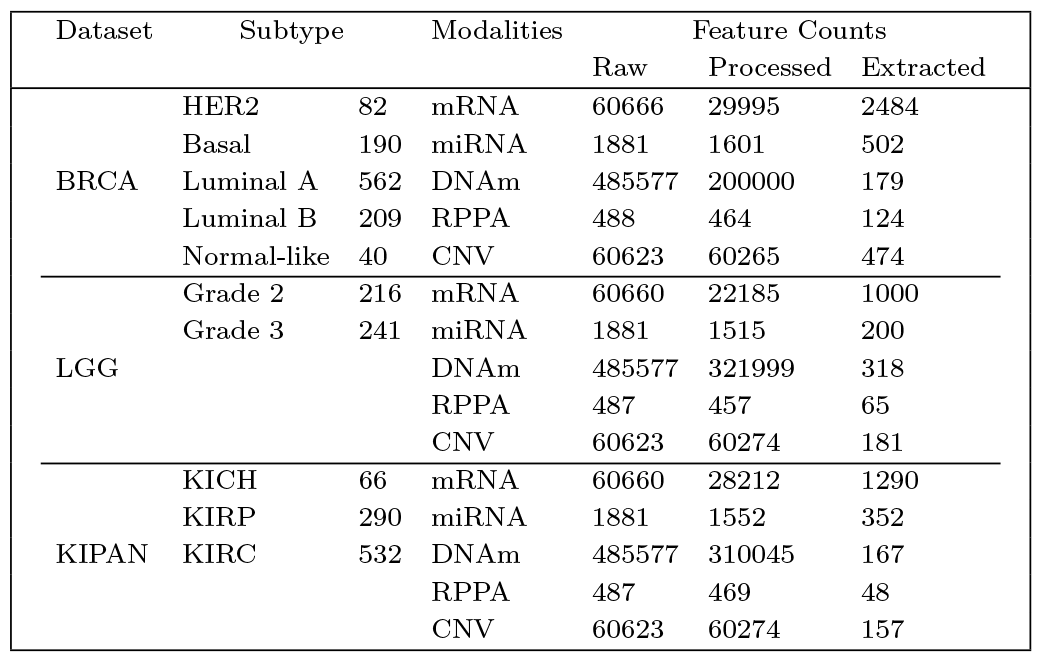
Summary of TCGA Datasets.

BRCA PAM50 is a 50-gene signature used to sub-type breast cancer into 5 classifications; Normal-like, Basal-like, HER2-enriched, Luminal A and Luminal B(Parker et al., 2009; Kensler et al., 2019). Patients included in this dataset have a mutation to their BRCA gene and therefore have a larger risk of developing breast cancer. Sub-typing by gene expression separates the carcinomas by varying biological properties and prognoses. For example, Luminal A has the best prognosis, while HER2 and Basal are considered more aggressive forms of cancer(Kensler et al., 2019). The KIPAN dataset consists of three categories separated by chromosomal differences (Tabibu et al., 2019). Clear Renal Cell Carcinoma (KIRC) is characterised by loss of chromosome 3p, Papillary Renal Cell Carcinoma (KIRP) is characterised by loss of chromosome 9p and Chromophobe Renal Cell Carcinoma (KICH) is characterised by loss of multiple other chromosomes. The LGG dataset consists of grade 2 and grade 3 which are characterised by the World Health Organisation based on their histopathologic characteristics (Forst et al., 2014). All of these datasets categorise a heterogeneous disease by a genetic association, making them suitable tasks for classification. They were chosen to demonstrate the generalisability of MOGDx to different diseases, as well as to benchmark the performance of MOGDx against other integrative methods (Wang et al., 2021; Li et al., 2022; Zhang et al., 2022).

### Performance & Evaluation

The performance of MOGDx was compared to existing network integrative methods that perform heterogeneous disease classification. Ablation experiments were also performed to understand the importance of different components of MOGDx. The performance metrics used to compare the classification performance of MOGDx were accuracy and F1-score (F1). The F1 score was calculated by the mean F1 score of each class, weighted by the size of that class. The F1 score was also weighted by the percentage of samples included in that analysis. The percentage of samples was calculated by the dividing the number of patients included in that analysis divided by the total number of patients available in that dataset. k-fold cross validation was performed with 5 randomly generated splits to obtain the mean and standard deviation metrics reported. Within each split, the training set was randomly split into training and validation sets to produce an overall train/validation/test split of 68%/12%/20% respectively.

#### MOGDx achieves superior performance when integrating a variable number of modalities while including a larger number of samples

The classification performance of MOGDx was compared to similar PSN multi-omic integrative methods as well as benchmark classification algorithms namely; Support Vector Machine (SVM), L1 regularized linear regression (Lasso) and gradient tree boosting (XGBoost). Table 2 shows MOGDx outperforms all benchmark classification algorithms. This demonstrates the predictive power of integrating multiple omic measures in these tasks.

**Table 2.** Summary of Results.

| Method | Dataset | Number of Modalities | Number of Samples | Number of Classes | Accuracy | F1 |
| --- | --- | --- | --- | --- | --- | --- |
| MOGDx | BRCA | 4 | 1083 | 5 | $0.893 \pm 0.014$ | $0.874 \pm 0.012$ |
| | BRCA | 4 | 1043 | 4 | $0.904 \pm 0.014$ | $0.887 \pm 0.016$ |
| | LGG | 1 | 457 | 2 | $0.899 \pm 0.016$ | $0.881 \pm 0.019$ |
| | KIPAN | 4 | 888 | 3 | $0.958 \pm 0.003$ | $0.948 \pm 0.004$ |
| MOGONET | BRCA | 3 | 875 | 5 | $0.829 \pm 0.018$ | $0.825 \pm 0.016$ |
| | LGG | 3 | 510 | 2 | $0.816 \pm 0.016$ | $0.814 \pm 0.014$ |
| | KIPAN | 3 | 658 | 3 | $0.999 \pm 0.002$ | $0.999 \pm 0.002$ |
| MoGCN | BRCA | 3 | 511 | 4 | $0.898 \pm 0.025$ | $0.902 \pm 0.024$ |
| | KIPAN | 3 | 698 | 3 | $0.977 \pm 0.017$ | $0.977 \pm 0.017$ |
| SVM | BRCA | 1 | 869 | 5 | $0.782 \pm 0.033$ | $0.721 \pm 0.030$ |
| Lasso | BRCA | 1 | 1047 | 5 | $0.829 \pm 0.014$ | $0.771 \pm 0.012$ |
| XGBoost | BRCA | 1 | 1047 | 5 | $0.762 \pm 0.036$ | $0.692 \pm 0.033$ |

The optimal MOGDx performance is shown for each dataset. 4 of 5 available modalities were used for both BRCA and KIPAN. Only DNAm was used on the LGG dataset as it achieved the best accuracy while still including maximum number of samples. The performance reported by MOGONET(Wang et al., 2021) was achieved using mRNA, miRNA and DNAm. The performance reported by MoGCN(Li et al., 2022) was achieved using CNV, mRNA and RPPA. The performances reported on SVM, Lasso and XGBoost methods were achieved using the omic measure which gave the highest accuracy.

MOGDx outperforms comparative integrative methods, MOGONET (Wang et al., 2021) and MoGCN (Li et al., 2022). On the BRCA dataset, MOGDx identified a combination of mRNA, DNAm, CNV and RPPA as the optimal integration of modalities. It performs better in both accuracy and F1 metrics compared to MOGONET. It achieves comparable performance to MoGCn when trained on four classes, and crucially retains double the number of samples. In supplementary figure S5, it can be seen that the Normal-like class is a difficult sub-type to classify. This is due to the small number of samples and the likelihood of these samples to go on to develop into one of the other sub-types. MoGCN does not include this sub-type in their classification, simplifying the task, resulting in higher accuracy. Interestingly, MOGDx strongly associates some of the Normal-like samples with other sub-types. This could suggest predictive power of this method to identify early signatures of BRCA. MOGDx identified a single omic measure, DNAm, which achieved optimal performance on the LGG dataset. All labelled samples were available in this single omic measure. While MOGDx did significantly outperform MOGONET on this dataset, there is a relatively large difference in number of samples. MOGONET obtained their data from Broad GDAC Firehose which stores TCGA data version from 2016 which could explain this discrepancy. Finally, MOGDx achieves slightly lower metrics on the KIPAN dataset compared to MOGONET and MoGCN. It identified miRNA, CNV, DNAm and RPPA as the optimal combination of modalities for integration. Once again, the number of samples differ due to differences between methods and differences in data collection. The lower performance of MOGDx in this dataset could be due to the imputation methods applied to account for missing samples in one or more data modality.

MOGDx can incorporate greater number of samples and modalities in its methodology. MOGONET and MoGCN are limited to the intersection of samples, which reduce the number of samples included in their analysis when more modalities are included. This is evident as both Lasso and XGBoost have greater number of samples available when trained on mRNA only. Conversely, MOGDx can incorporate all available samples due to imputation methods employed without a significant degradation in performance. Moreover, MOGONET and MoGCN are fixed to the integration of three modalities. The flexibility of MOGDx allows any number of modalities to be included, resulting in significantly improved performance on the LGG dataset as per Table 2. This flexibility also provides insight into the information content of each modality. MOGDx, facilitates the analysis of individual as well as combined omics. This can be utilised to identify the best combination of modalities and the added information content of including additional modalities.

#### The performance of MOGDx varies under different omic data types for different classification tasks

Figure 2.A shows the performance of MOGDx varies significantly when different numbers of omic measures are integrated. There exists a trade-off between modality integration and performance. As can be seen in supplementary figure S4, typically three to four modalities are required to ensure full sample coverage. Figure S2 shows that some omic measures are significantly more predictive than others. We can see that BRCA and KIPAN benefit from an integrated approach, while LGG is most easily predicted using only DNAm. Furthermore, we can see that for KIPAN that mRNA is not included in the least predictive nor, the most predictive. In fact, some omics, which are less predictive individually, are more predictive than mRNA when integrated. This suggests these omics are capturing different sources of variation, highlighting the importance of a multi-omic approach. Overall, omic measures should be included if they improve performance or contribute a significant number of samples not contained in other measures. In order to test this, all combinations of modalities were trained using MOGDx. The modality or combination of modalities which achieved the best classification performance and including all samples were reported in Table 2, with the performance of all other combinations reported in supplementary tables S2-S4. In the BRCA and KIPAN datasets, integration of 4 modalities resulted in optimal performance. mRNA, DNAm, CNV and RPPA were most predictive for BRCA. miRNA, DNAm, CNV and DNAm were most predictive for KIPAN. Although including more modalities does not always produce the optimal accuracy, it does reduce the standard deviation of accuracy estimates from cross-validation, as per Figure 2. Conversely, training MOGDx on only DNAm resulted in the best performance on the LGG dataset. The DNAm dataset had all samples present, as per Figure S2, meaning there was full sample coverage. In this case, it is clear from Figure 2 that DNAm was the only informative modality for tumour grade in the LGG dataset, and including more modalities increased the standard deviation of accuracy estimates without increasing overall accuracy or samples. This demonstrates that flexibility to train on any number and/or combination of modalities is an important requirement for integrative network approaches.

**Fig. 2:**
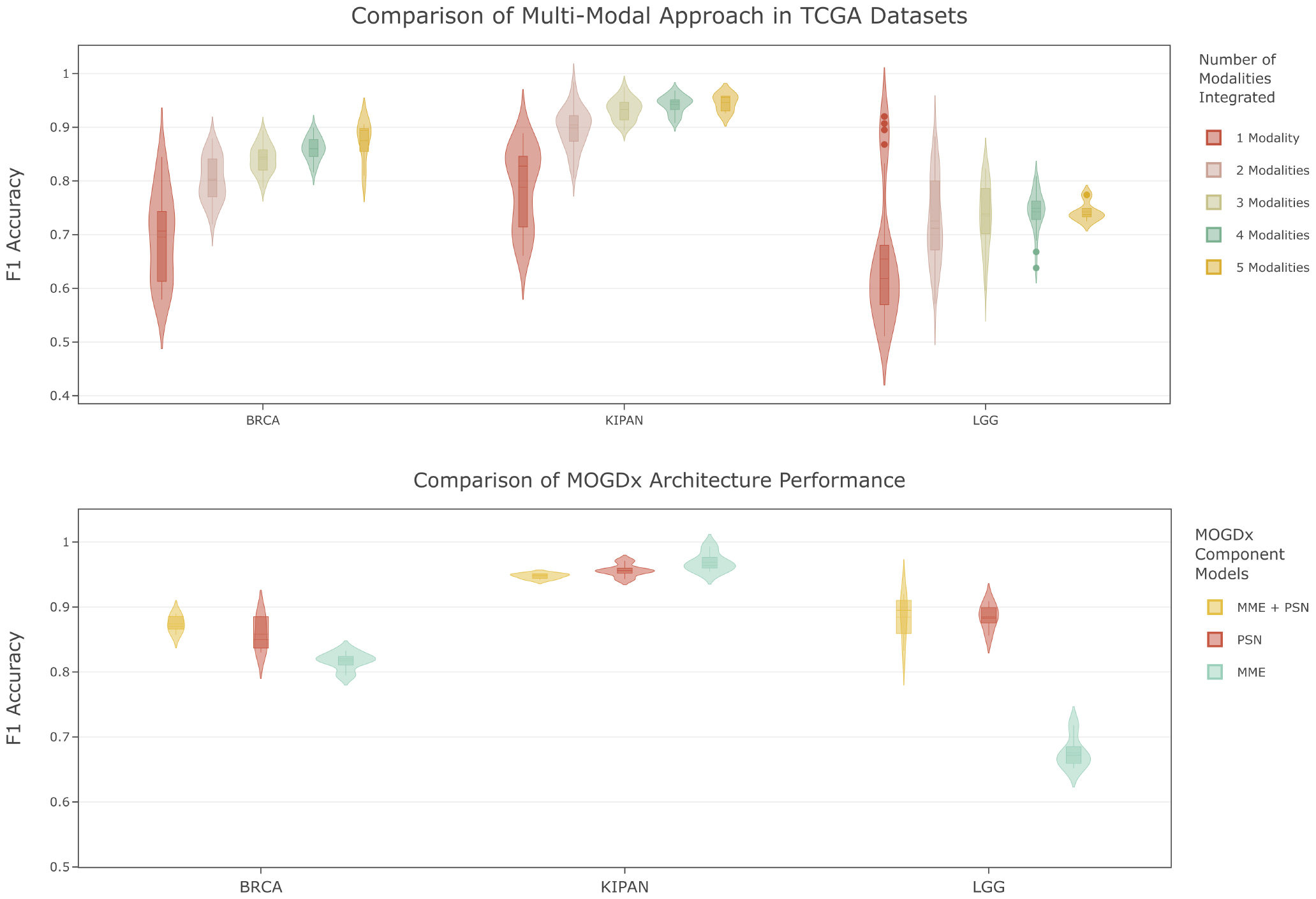
A Modality Importance. F1 accuracy per number of integrated modalities across the three datasets. The distribution of the violin plots were generated using the F1 accuracy across all folds for the optimal hyperparameters of each modality’s model. **B PSN Importance** — Performance of MOGDx on the best model when trained on MME only, PSN only and when trained on MME + PSN (MOGDx). The MME model has been trained on the MME only by removing all edges from the PSN. Similarly, the PSN model was trained by one hot encoding each node feature, thus only allowing the GCN to learn from the structure of the PSN. MOGDx learns from both MME node features and PSN.

#### Optimal performance is achieved when MOGDx is trained on fused PSN and node features

Figure 2.B demonstrates the combined predictive power of the MME and the PSN. Tables S5-S7 show the accuracy and F1 results of the different component models for each cross validation fold. For both the BRCA and LGG datasets, the best mean F1 accuracy is achieved when using the combined MME and PSN architecture. The MME achieves the highest mean F1 accuracy in KIPAN, however it achieves significantly poorer performance in LGG and has higher variation in its train/test splits. The predictive power from learning relationships between patient samples is strong and consistent, but is susceptible to a larger amount of variation due to train/test splits compared to the combined components in the BRCA and KIPAN datasets. Integrating the PSN with the MME reduces the variation in accuracy of the GCN to splits in the data. It is known that GNN’s are sensitive to train/test and validation splits. Shchur et al. (2019) showed that classification metrics are susceptible to inflated results when models are trained on the same splits. To overcome this limitation of GNN’s MOGDx was trained on shuffled splits of the data. Embedding a joint reduced representation of all modalities as node features allows the GNN to learn from more than just the network structure, reducing this variability furthermore, as can be seen in Figure 2.B. In this manner, MOGDx achieves a balanced trade-off between accuracy and robustness by integrating the MME with the PSN.

#### MOGDx can identify relevant biological markers of heterogeneous diseases

Ablation experiments were performed to determine which omic measures were most predictive of each classification task, with the results shown in Tables S2-S4. It was determined that mRNA and DNAm are highly predictive of BRCA and KIPAN, while only DNAm was predictive of LGG. mRNA and DNAm have established enrichment pipelines facilitating this analysis, thus these modalities were analysed for relevant biological markers in all datasets. Enrichment analysis was performed on the extracted features from the differential expression analysis on mRNA and penalised regression analysis on DNAm modalities in all datasets.

Table 3 shows the functional processes associated with mRNA and DNAm respectively for; BRCA PAM50 sub-type classification, KIPAN sub-type classification and LGG grade classification. Two gene sets were analysed, KEGG and MSigDB, for functional and biological enrichment processes and the full results of the enrichment analyses are given in tables S8 and S9.

**Table 3.** Summary of Enriched mRNA and DNAm Pathways in TCGA Dataset.

| Dataset | Modality | Gene Set |  |
| --- | --- | --- | --- |
|  |  | KEGG 2021 Human | MSigDB Hallmark 2020 |
| BRCA | mRNA | Pentose and glucuronate interconversions | G2-M Checkpoint |
|  |  | Cell cycle | Androgen Response |
|  |  | Homologous recombination | Myc Targets V1 |
|  | DNAm | Axon guidance | Estrogen Response Early |
|  |  | Proteoglycans in cancer | Epithelial Mesenchymal Transition |
|  |  | Pathways in cancer | Myogenesis |
| LGG | mRNA | Nicotine addiction | G2-M Checkpoint |
|  |  | Systemic lupus erythematosus | E2F Targets |
|  |  | Type I diabetes mellitus | Epithelial Mesenchymal Transition |
|  | DNAm | Axon guidance | Epithelial Mesenchymal Transition |
|  |  | Proteoglycans in cancer | Estrogen Response Early |
|  |  | Focal adhesion | Apical Junction |
| KIPAN | mRNA | Oxidative phosphorylation | Oxidative Phosphorylation |
|  |  | Pentose and glucuronate interconversions | Allograft Rejection |
|  |  | Intestinal immune network for IgA production | Epithelial Mesenchymal Transition |
|  | DNAm | Axon guidance | Estrogen Response Early |
|  |  | Human papillomavirus infection | Myogenesis |
|  |  | Proteoglycans in cancer | Epithelial Mesenchymal Transition |

The functional processes found to be enriched in the BRCA mRNA dataset in both gene sets are related to the life cycle of a cell, with good concordance between the two gene sets and corroborated with established research (Yarden et al., 2002; Deng, 2006). Again, the DNAm enriched functional processes show good concordance between KEGG and MSigDB gene sets in BRCA. These enriched processes have established involvement with the development and growth of cancer tumours (Tzanakakis et al., 2020). The enriched DNAm processes in BRCA overlap significantly with LGG and KIPAN. This finding is unsurprising considering we have assessed three cancer tumour datasets. It does, however, highlight the ability of MOGDx to consistently identify relevant pathways, as well as motivating the use of DNAm to differentiate between different cancers and cancer subtypes. The concordance between enriched pathways in mRNA and DNAm differs more in BRCA than it does in LGG and KIPAN. This suggests that the enriched features of mRNA and DNAm are more orthogonal in BRCA, which could explain the inclusion of both of these modalities in the best performing model for this dataset. Conversely, in the LGG and KIPAN datasets, we see more overlap, with Epithelial Mesenchymal Transition enriched in both mRNA and DNAm. This indicates that similar information is captured in both these omics and thus, justifies the exclusion of one of them from the optimal modality combination. Given, mRNA was less predictive in both LGG and KIPAN, shown in Tables S2 and S3, and the apparent overlap in information, it is less surprising that this modality was excluded from the optimal combination of modalities in these datasets.

## Discussion

Disease heterogeneity has moved medical research from a population-based perspective towards a personalised approach where diagnosis, prognosis and treatments are selected based on biomedical characteristics. Driving this movement is the development of large, diverse omic technologies and studies which provide labelled biomedical data at unprecedented levels. The integration of these omic measures offer the opportunity to build quantitative models, which can aid the understanding of heterogeneous disease architectures and inform clinical guidance. Therefore, a tool which can flexibly incorporate omic measures and identify specific biomedical characteristics based on these labels has the ability to redefine heterogeneous diseases.

We propose MOGDx, a network integrative architecture for the classification of heterogeneous diseases. What separates MOGDx from its competitors is the flexibility in its integration of omic measures, its inclusion of all available patient samples and its leverage of the predictive power of PSN‘s. MOGDx includes omic measures, which either improves predictive performance or include patients who may only have samples in one omic measure. This allows users to fine tune to the most predictive modalities while incorporating the maximum number of patient samples in an analysis. In this analysis, we maximised data usage while maintaining competitive or state-of-the-art performance on a variety of classification tasks. Fundamental to the predictive performance of MOGDx is the integration of PSN’s. In this analysis, we have shown that patient similarity is a very effective determinant of heterogeneous disease sub-typing and grading. The use of PSN’s is analogous to clinical diagnosis, where a diagnostician will compare a new patient to a database of similar cases. Similarly, MOGDx captures the variability in similarity and uses this to perform accurate sub-type classification and grading.

The application of MOGDx has been benchmarked on three cancer datasets from the TCGA, namely; BRCA, LGG and KIPAN. Cancer is widely regarded as a highly heterogeneous disease however, MOGDx was able to accurately classify breast cancer sub-types, kidney cancer sub-types and brain tumour grades from integrated omic data. MOGDx identified the optimal combination of modalities which resulted in greater patient coverage while maintaining a state-of-the-art classification performance compared to its competitors, as per Table 2.

Interpretability is an important aspect to consider for biomedical applications in order to transform research into novel diagnoses, grades or treatments. We have demonstrated the interpretability of MOGDx in several ways. Firstly, through leave one out experiments we have identified the modalities which are most predictive of the classification task, their most important features, and some enriched functional pathways as summarised in Table 3. Through enrichment analyses on the extracted features in the mRNA and DNAm modalities we have identified the main drivers of their variability and combining this with individual omic prediction accuracy yields novel insights and makes MOGDx more explainable.

The use of different omics modalities allows us to assay different parts of the biological systems involved in disease mechanism, their integration can help reduce biological noise improving signal and allowing for the identification of previously undetectable informative features. Understanding which omic modalities are most predictive for a given disease can allow us to design more efficient and informative experiments, minimising impacts on patients and reducing costs. Further, because different omics modalities capture different components of the genetic and environmental contributions to disease their integration can help us to gain a more complete picture of disease. We performed enrichment analysis on mRNA and DNAm modalities in all TCGA datasets. MOGDx was able to identify features enriched in processes and genes relating to the pathology and prognosis of the disease. These findings were supported by similar findings in the literature demonstrating MOGDx’s ability to identify important omic markers. We have demonstrated in this work that the MOGDx architecture can successfully produce interpretable and reproducible insights into multiple heterogeneous diseases.

The main drawback of MOGDx is that the GNN algorithm employed in this analysis, is a transductive algorithm. Transductive algorithms require the entire network to be available during training. In a clinical setting, this is not possible as it will be required to perform predictions on unseen patients. Hence, it will be required to extend MOGDx to an inductive algorithm which will not require the entire network to be available during training and can make predictions on unseen patients. In summary, MOGDx is a flexible and accurate classification tool which can be applied to a broad range of heterogeneous diseases.

## Conclusion

In this paper, we present MOGDx, a command line tool for the integration of omic datasets to perform classification tasks for biomedical applications. We show state-of-the-art or comparative performance on three cancer datasets. We highlight the importance of flexibility when integrating omic data for classification tasks and show the predictive power of representing data as PSN’s. The advantages of MOGDx are that we can maintain state-of-the-art classification accuracy while including a significantly larger number of patients. It also offers layers of interpretability. It can integrate any number of modalities, providing insight into the informative content of modalities, such as which is the most informative modality and if integrating additional modalities increases the predictive power. Furthermore, combining this with enrichment analysis has shown robust performance of MOGDx, and possible explanatory behaviour when selecting the optimal combination of modalities.

## Methods

### Framework of MOGDx

The framework for MOGDx can be summarised into three main components; 1) Pre-processing and feature extraction, 2) Graph generation and SNF, 3) Graph Convolutional Network with Mulit-Modal Encoder (GCN-MME) training and classification. Before integration, each modality is treated individually. An individual modality will undergo processing steps where an expression matrix and meta file for each modality is created. Feature extraction will be performed on this expression matrix and a PSN generated on the most important features. This PSN will then be used to create a network based on the KNN algorithm. Integration is performed using SNF on the networks and the fused network along with each expression matrix are inputted into the GCN-MME. The MME encodes each expression matrix and decodes the reduced representation to a shared decoding space. The nodes of the fused PSN are augmented with a vector representation from the shared embedding space of the MME. The GNN is trained in combination with the MME on the joined PSN augmented with MME shared embeddings for heterogeneous disease classifications. MOGDx is a command line tool which can integrate a variable number of omic measures. Specific details of each component are described in the following sections.

### Pre-processing and Feature Extraction

Pre-processing is performed to remove unwanted noise and variations in the data due to experimental or technical effects. For mRNA expression (mRNA) and micro RNA expression (miRNA) all genes which had either zero expression or zero variance in all samples were removed. Next, any samples which were more than 2 standard deviations from the mean node connectivity distance were removed. Differential expression was performed using a one-vs-the-rest methodology, and the most significantly differentially expressed genes were extracted.

The DNA methylation (DNAm) data downloaded from TCGA-Biolinks used multiple generations of Illumina Infinium DNA methylation arrays, however, they have been corrected and standardised using the SeSAMe(Zhou et al., 2018) pipeline. Further steps were taken to remove any CpG sites which contained missing values. Important CpG sites were identified by performing a penalised Logistic Regression algorithm and keeping any CpG sites which had a non-zero weight.

To overcome significant missingness in the Copy Number Variation (CNV) and Reverse Phase Protein Array (RPPA) datasets, sites which contained more than 50% missingness were removed, and mean imputation was performed. The CNV data was log transformed to give it a close to normal distribution, and penalised Logistic Regression was applied to both. The CNV and RPPA sites which had a non-zero weight were extracted.

### Graph Generation and Similarity Network Fusion

The modalities were represented as graphs and Similarity Network Fusion (SNF) was performed to integrate the modalities. A patient similarity matrix was first created for each modality. The Pearson correlation coefficient (Eq. 1) between the extracted features was used as a measure of similarity.

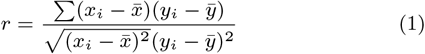

The K-Nearest Neighbours (KNN) algorithm was used to build the graph with edges created between the 15 nearest neighbours. SNF(Wang et al., 2014) was applied to fuse the graphs into a single network representing the full spectrum of the underlying data. SNF allows complimentary information to be shared between modalities, and also is effective in identifying novel relationships between patients. It also integrates missing patient samples inherently by complimenting a missing edge in one modality with the same relationship from others.

### Graph Convolutional Network - Multi-Modal Encoder Training and Classification

The architecture of the GCN-MME consists of two main parts; a Mulit-Modal Encoder (MME) and a Graph Convolutional Network (GCN), with the architecture shown in Figure S1. The MME consists of two linear layers. First, each modality is encoded to a reduced linear layer of dimension 500. Batch normalisation is performed and the output of this linear layer is encoded further to a second linear layer of arbitrary dimension. The motivation for using two layers is to capture interactions between features, as well as simply compressing the dimension of each modality. The output dimension of the encoder is specific to each modality and found by performing a hyperparameter search. Further batch normalisation and median imputation of missing patient samples on this encoded layer are performed. The encoded output of each modality is decomposed into the shared embedding space through mean pooling.

Each node in the fused PSN is augmented with the corresponding vector in the MME shared embedding space. The weights of the linear layers are calculated by performing back propagation through the GCN and MME to form a single fully supervised pipeline.

GNN’s are a powerful architecture for the learning of graph structure and information in a supervised setting. We implemented a GCN model from the Deep Graph Library in Python with a PyTorch backend. The differentiation between GCN and neural network architectures is their ability to learn from the local neighbourhood as opposed to handcrafted features. The performance of GCN and other GNN architectures has been demonstrated on a variety of benchmark tasks, hence extending their application to a biomedical setting is an exciting avenue with great potential.

GCN requires two inputs. A network, consisting of nodes and edges, and a vector of features for each node. For MOGDx, the network created was a PSN and the vector of features was a reduced feature representation from the MME. Formulating the GCN algorithm as a network represented by an adjacency matrix *A* ∈ *R*^*nxn*^ and a feature matrix *X* ∈ *R*^*nxd*^ where n is the number of patients and d is the latent dimension selected for the MME. The GCN then consists of stacked convolutional layers defined by Eq. 3(Kipf and Welling, 2017).

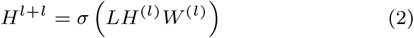

Where 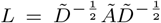 is the normalised graph Laplacian; Ã = *A* + *I* is the adjacency matrix; 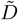 is the degree matrix of Ã *W* is the weight matrix learned during training; *σ* is the non-linear activation function, ELU activation in this case, *H*^(*l*)^ is the input to each layer and *H*^(0)^ corresponds to *X* the node feature matrix.

### Interpretability of MOGDx

Interpretability in biomedical applications is important to understand how specific features contribute to prediction so that therapeutic interventions or novel diagnoses can be well understood. MOGDx shows interpretability in a number of ways. Firstly, through ablation experiments, we can identify which omic measures are most predictive of the targeted outcome. Ablation experiments are widely adopted for feature importance and ranking in neural networks(Setiono and Liu, 1997). Similarly, we can treat entire modalities as features in MOGDx to identify the most informative. Enrichment analysis is a well understood methodology to map selected genes to their biological and molecular pathways. Functional enrichment analysis was carried out using the Gene Set Enrichment Analysis (GSEA) algorithm by Shi and Walker (2007) in Python on the extracted features from the mRNA datasets. Similarly, enrichment analysis was carried out using the mCSEA(Martorell-Marugán et al., 2019) algorithm in R on the extracted CpG sites from the DNAm datasets. Results from these analyses were further compared to existing literature. Receiver Operator Characteristic and Area Under Curve plots are shown in Figures S5-S7. Through these visualisations, we can assess which classes are most difficult to predict and obtain metrics for the overall accuracy of the model.

## Author contributions statement

BR gathered all data, performed analysis, designed the study, conducted experiments and drafted the manuscript. TIS contributed to analysis, results and discussions. TIS and REM supervised the study, revised the manuscript and approved the final version of the manuscript.

## Acknowledgments

This work was supported by the United Kingdom Research and Innovation [grant EP/S02431X/1], UKRI Centre for Doctoral Training in Biomedical AI at the University of Edinburgh, School of Informatics. For the purpose of open access, the author has applied a creative commons’ attribution [CC BY] licence to any author accepted manuscript version arising.

The results published here are in whole or part based upon data generated by the TCGA Research Network: https://www.cancer.gov/tcga

## Additional information

### Data Availability

All data is available to download from The Cancer Genome Atlas (TCGA)(https://www.cancer.gov/tcga). A script to download and obtain data exactly as it is presented is available from https://github.com/Barry8197/MOGDx.

### Supplementary Material

Supplementary Material is available in the accompanying files S1FiguresTablesMOGDx.pdf and S2ExtractedFeaturesMOGDx.pdf

**Author Name**. This is sample author biography text. The values provided in the optional argument are meant for sample purposes. There is no need to include the width and height of an image in the optional argument for live articles. This is sample author biography text this is sample author biography text this is sample author biography text this is sample author biography text this is sample author biography text this is sample author biography text this is sample author biography text this is sample author biography text.

**Author Name**. This is sample author biography text this is sample author biography text this is sample author biography text this is sample author biography text this is sample author biography text this is sample author biography text this is sample author biography text this is sample author biography text.

© The Author 2022. Published by Oxford University Press. All rights reserved. For permissions, please **1**

