## Supplementary Figures and Tables for "Multi-Omic Graph Diagnosis (MOGDx) : A data integration tool to perform classification tasks for heterogeneous diseases"

*Journal Title Here*, 2022, 1–1
doi: DOI HERE
Advance Access Publication Date: Day Month Year
Paper

### Multi-Omic Graph Diagnosis (MOGDx) : A data integration tool to perform classification tasks for heterogeneous diseases

**Barry Ryan,[1,*] Riccardo E. Marioni[2] and T. Ian Simpson[1]**

[1]School of Informatics, University of Edinburgh, 10 Crichton Street, EH8 9AB, Edinburgh, UK and [2]Centre for Genomic and Experimental Medicine, Institute of Genetics and Cancer , University of Edinburgh, Crewe Rd S, EH4 2XU, Edinburgh, UK

FOR PUBLISHER ONLY Received on Date Month Year; revised on Date Month Year; accepted on Date Month Year

#### Supplementary Figures

#### Supplementary Tables

© The Author 2022. Published by Oxford University Press. All rights reserved. For permissions, please 

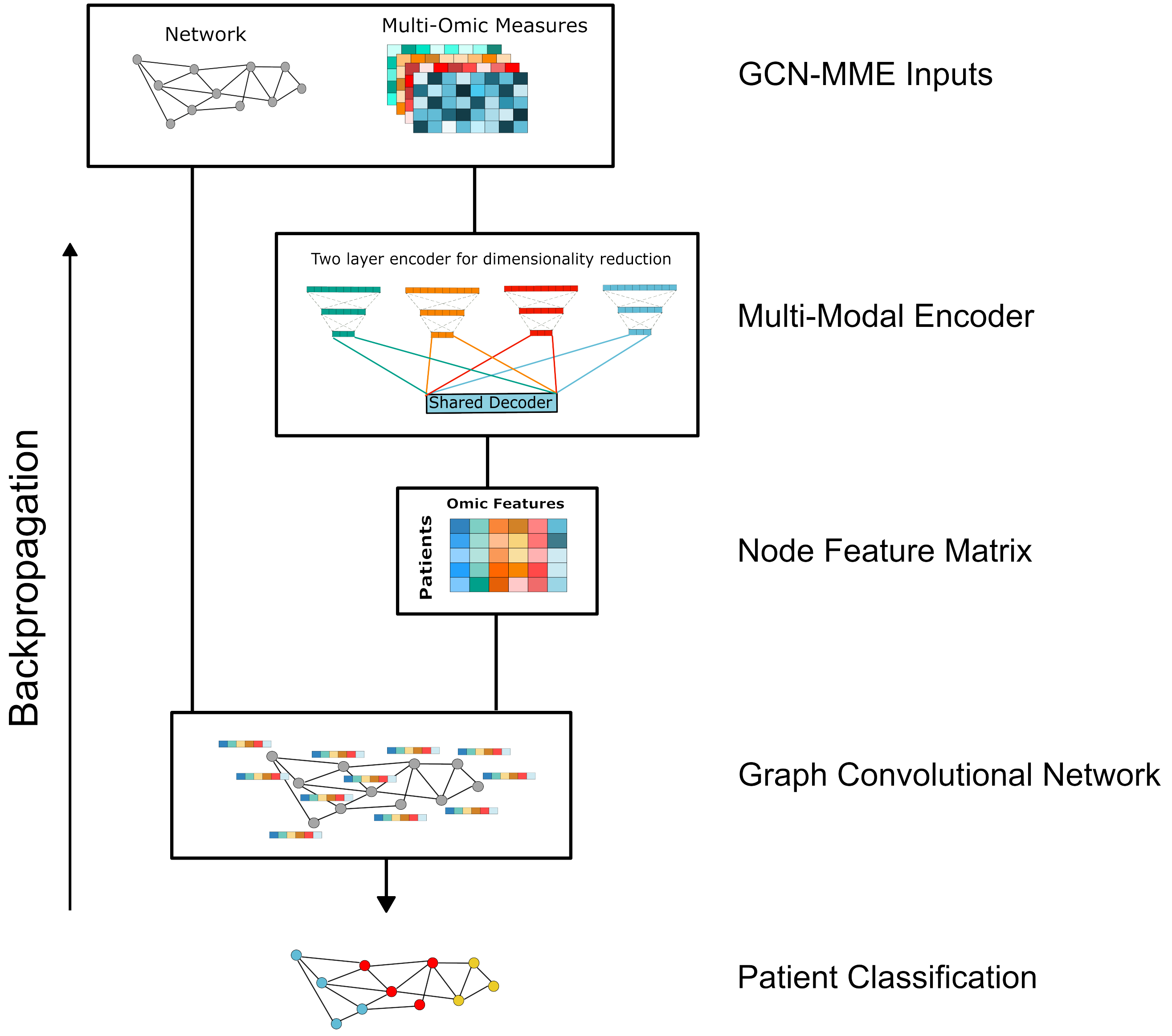


Fig. 1: Graph Convolutional Network - Multi-Modal Encoder Architecture

**Table 1.** Model Parameters for Optimal Performance on Each Dataset

| Dataset Model | MME Modality Reduced Dimension | | | | | Shared Decoder Dimension | GCN Dimension |
|---|---|---|---|---|---|---|---|
| | mRNA | miRNA | DNAm | CNV | RPPA | | |
| BRCA | 32 | X | 32 | 16 | 16 | 32 | 32 |
| LGG | X | X | 32 | X | X | 32 | 32 |
| KIPAN | X | 16 | 32 | 16 | 16 | 64 | 128 |

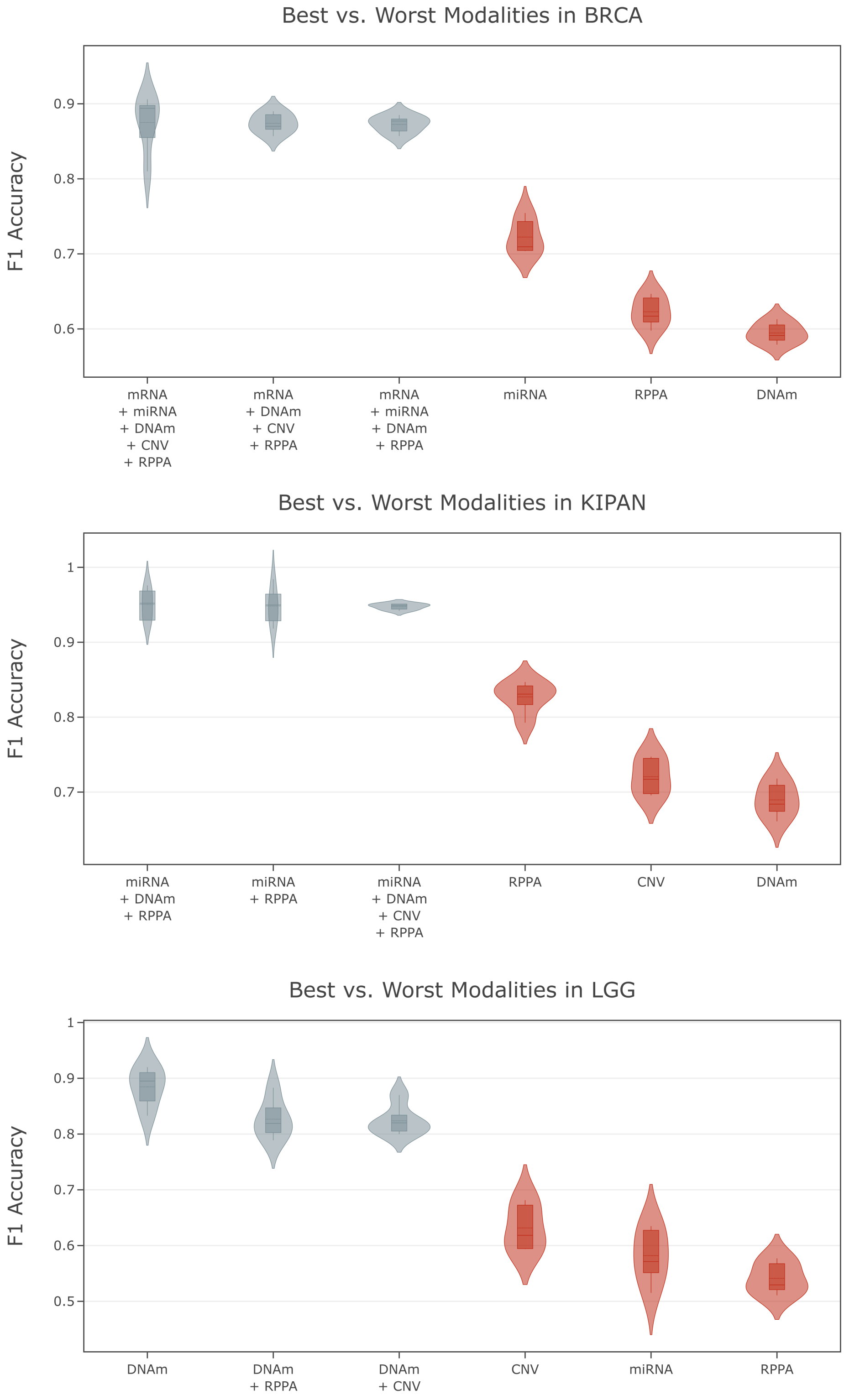


Fig. 2: Best and Worst Performing Modalities in Each Dataset

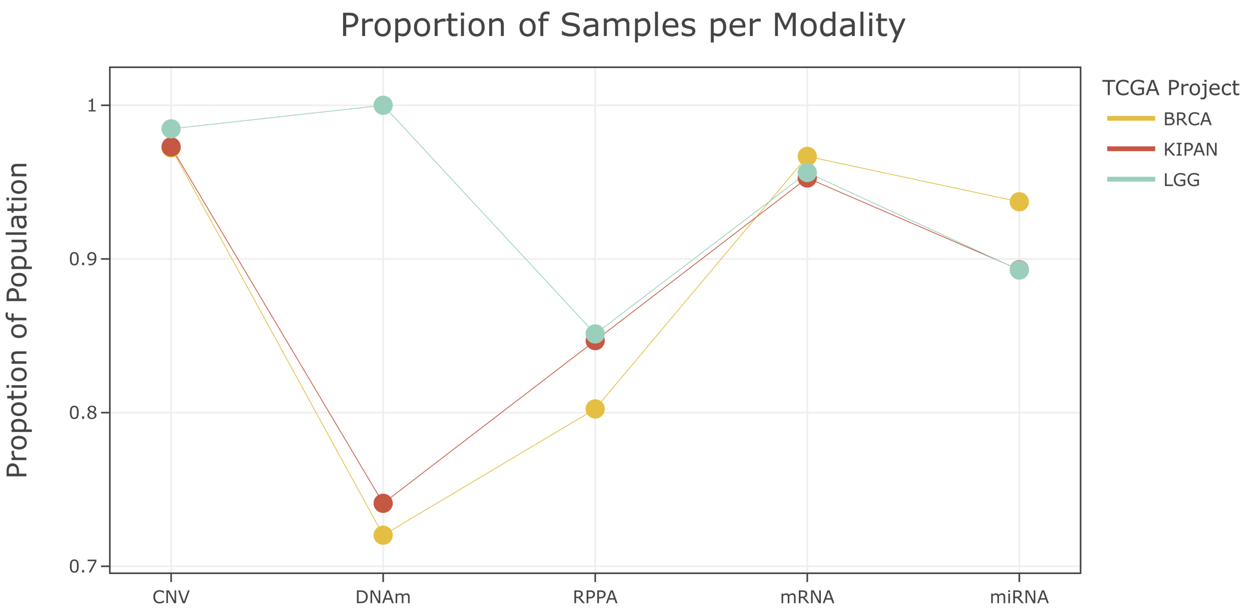


Fig. 3: Number of Samples per Modality in TCGA Datasets

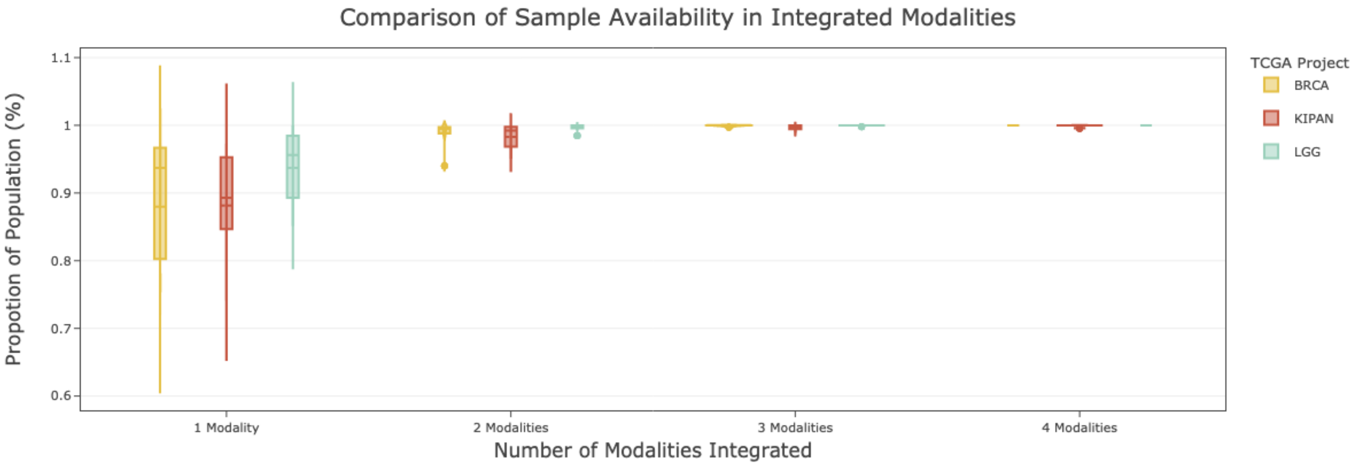


Fig. 4: Percentage of Samples Included when Integrating Modalities

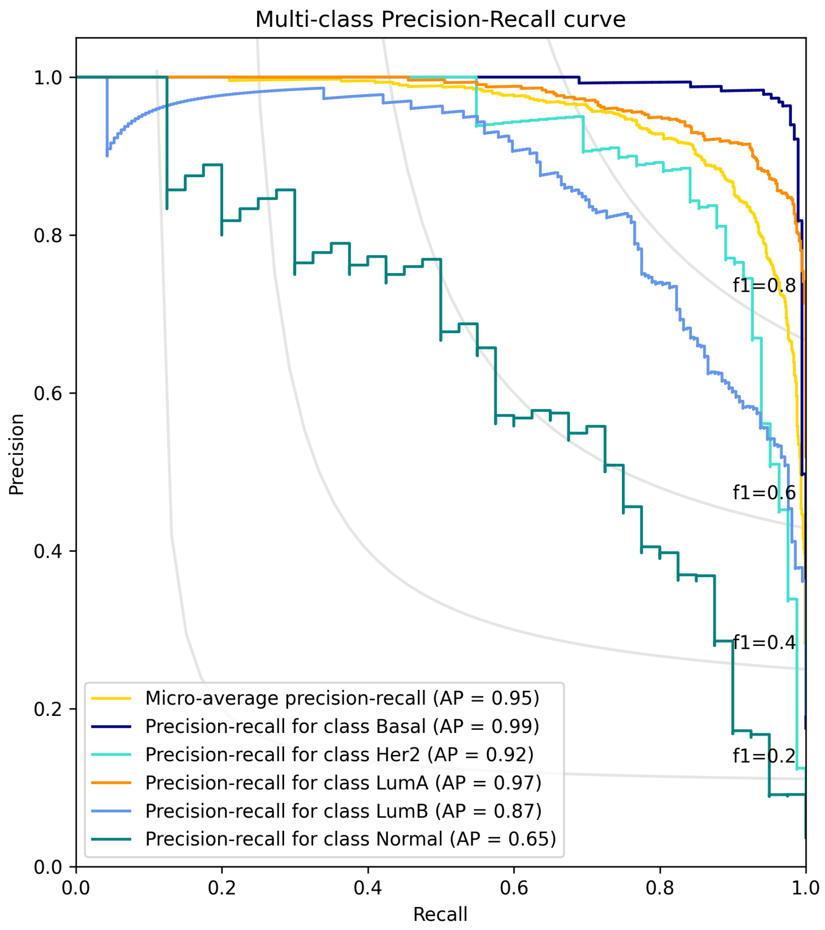


Fig. 5: BRCA Precision Recall Curve

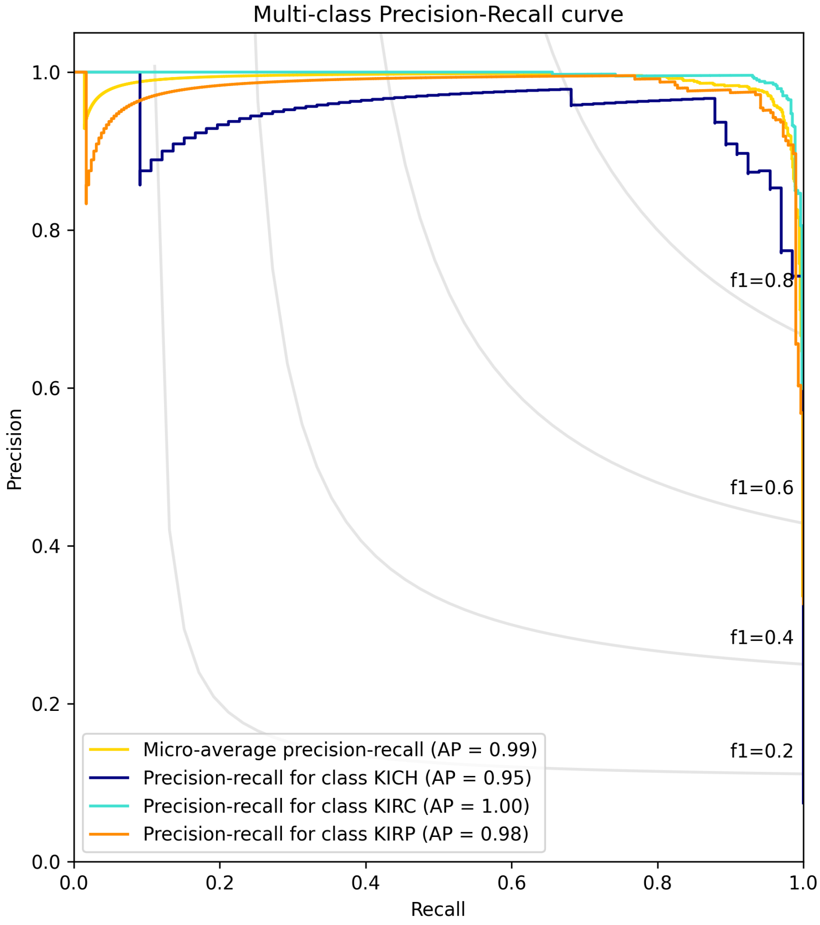


Fig. 6: KIPAN Precision Recall Curve

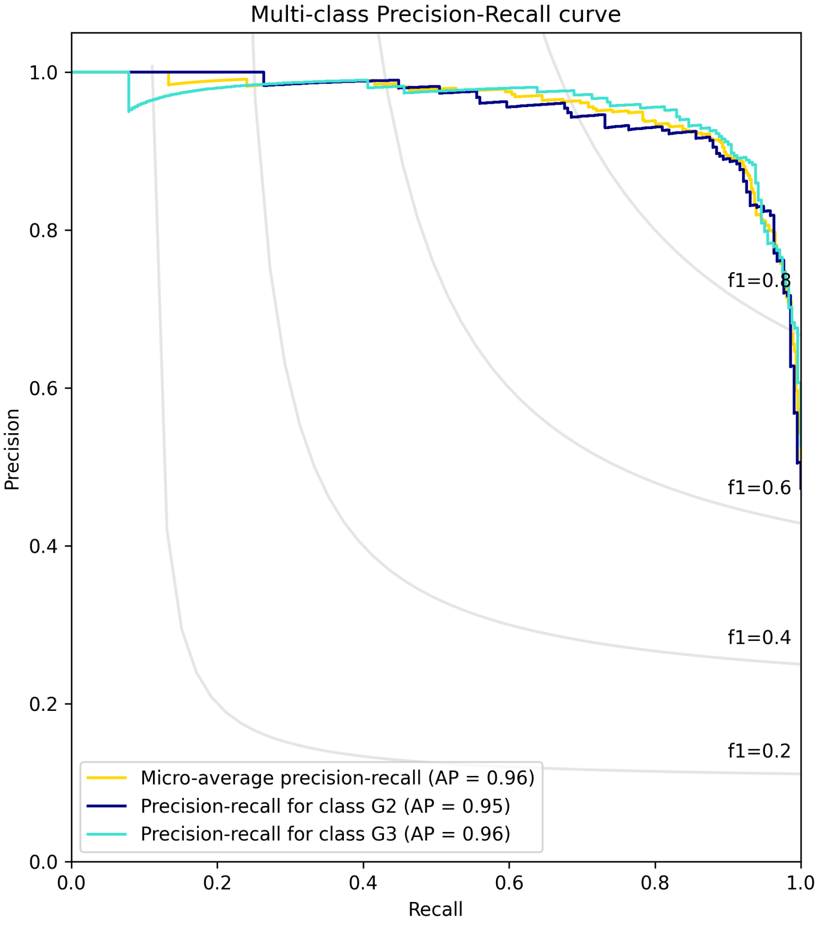


Fig. 7: LGG Precision Recall Curve

**Table 2.** KIPAN: Optimal Modality Performance Summary. These accuracies are not weighted by the percentage of sample availability.

| Project | Modality | Accuracy | F1 |
|---|---|---|---|
| KIPAN | CNV | 0.76 | 0.74 |
| | DNAm | 0.95 | 0.93 |
| | miRNA | 0.95 | 0.94 |
| | RPPA | 0.98 | 0.98 |
| | mRNA | 0.92 | 0.9 |
| | miRNA + DNAm | 0.95 | 0.94 |
| | miRNA + CNV | 0.89 | 0.87 |
| | mRNA + RPPA | 0.95 | 0.94 |
| | mRNA + DNAm | 0.94 | 0.92 |
| | mRNA + miRNA | 0.93 | 0.92 |
| | mRNA + CNV | 0.9 | 0.88 |
| | CNV + RPPA | 0.91 | 0.89 |
| | DNAm + CNV | 0.88 | 0.86 |
| | DNAm + RPPA | 0.97 | 0.96 |
| | miRNA + RPPA | 0.97 | 0.96 |
| | miRNA + DNAm + RPPA | 0.97 | 0.96 |
| | mRNA + miRNA + CNV | 0.93 | 0.92 |
| | mRNA + miRNA + DNAm | 0.94 | 0.93 |
| | mRNA + CNV + RPPA | 0.95 | 0.94 |
| | mRNA + miRNA + RPPA | 0.96 | 0.94 |
| | DNAm + CNV + RPPA | 0.95 | 0.94 |
| | miRNA + CNV + RPPA | 0.95 | 0.94 |
| | mRNA + DNAm + CNV | 0.94 | 0.92 |
| | miRNA + DNAm + CNV | 0.94 | 0.93 |
| | mRNA + DNAm + RPPA | 0.96 | 0.94 |
| | mRNA + miRNA + CNV + RPPA | 0.95 | 0.94 |
| | mRNA + miRNA + DNAm + CNV | 0.95 | 0.94 |
| | mRNA + miRNA + DNAm + RPPA | 0.96 | 0.95 |
| | mRNA + DNAm + CNV + RPPA | 0.96 | 0.94 |
| | miRNA + DNAm + CNV + RPPA | 0.96 | 0.95 |
| | mRNA + miRNA + DNAm + CNV + RPPA | 0.96 | 0.95 |

**Table 3.** LGG Optimal Modality Performance. These accuracies are not weighted by the percentage of sample availability.

| Project | Modality | Accuracy | F1 |
|---|---|---|---|
| LGG | CNV | 0.66 | 0.64 |
| | DNAm | 0.90 | 0.88 |
| | miRNA | 0.68 | 0.65 |
| | RPPA | 0.65 | 0.64 |
| | mRNA | 0.69 | 0.66 |
| | miRNA + DNAm | 0.84 | 0.82 |
| | miRNA + CNV | 0.68 | 0.66 |
| | mRNA + RPPA | 0.72 | 0.69 |
| | mRNA + DNAm | 0.77 | 0.74 |
| | mRNA + miRNA | 0.7 | 0.68 |
| | mRNA + CNV | 0.72 | 0.69 |
| | CNV + RPPA | 0.73 | 0.7 |
| | DNAm + CNV | 0.85 | 0.82 |
| | DNAm + RPPA | 0.85 | 0.83 |
| | miRNA + RPPA | 0.67 | 0.65 |
| | miRNA + DNAm + RPPA | 0.81 | 0.78 |
| | mRNA + miRNA + CNV | 0.72 | 0.7 |
| | mRNA + miRNA + DNAm | 0.75 | 0.72 |
| | mRNA + CNV + RPPA | 0.72 | 0.7 |
| | mRNA + miRNA + RPPA | 0.69 | 0.68 |
| | DNAm + CNV + RPPA | 0.82 | 0.79 |
| | miRNA + CNV + RPPA | 0.7 | 0.67 |
| | mRNA + DNAm + CNV | 0.8 | 0.77 |
| | miRNA + DNAm + CNV | 0.82 | 0.8 |
| | mRNA + DNAm + RPPA | 0.76 | 0.74 |
| | mRNA + miRNA + CNV + RPPA | 0.7 | 0.69 |
| | mRNA + miRNA + DNAm + CNV | 0.79 | 0.76 |
| | mRNA + miRNA + DNAm + RPPA | 0.78 | 0.75 |
| | mRNA + DNAm + CNV + RPPA | 0.79 | 0.76 |
| | miRNA + DNAm + CNV + RPPA | 0.79 | 0.76 |
| | mRNA + miRNA + DNAm + CNV + RPPA | 0.77 | 0.74 |

**Table 4.** BRCA Optimal Modality Performance. These accuracies are not weighted by the percentage of sample availability.

| Project | Modality | Accuracy | F1 |
|---|---|---|---|
| BRCA | CNV | 0.67 | 0.75 |
| | DNAm | 0.84 | 0.83 |
| | miRNA | 0.77 | 0.77 |
| | RPPA | 0.8 | 0.78 |
| | mRNA | 0.87 | 0.84 |
| | miRNA + DNAm | 0.82 | 0.81 |
| | miRNA + CNV | 0.78 | 0.77 |
| | mRNA + RPPA | 0.87 | 0.84 |
| | mRNA + DNAm | 0.88 | 0.86 |
| | mRNA + miRNA | 0.86 | 0.84 |
| | mRNA + CNV | 0.87 | 0.84 |
| | CNV + RPPA | 0.79 | 0.79 |
| | DNAm + CNV | 0.78 | 0.77 |
| | DNAm + RPPA | 0.83 | 0.82 |
| | miRNA + RPPA | 0.81 | 0.8 |
| | miRNA + DNAm + RPPA | 0.85 | 0.84 |
| | mRNA + miRNA + CNV | 0.87 | 0.85 |
| | mRNA + miRNA + DNAm | 0.88 | 0.86 |
| | mRNA + CNV + RPPA | 0.87 | 0.85 |
| | mRNA + miRNA + RPPA | 0.87 | 0.85 |
| | DNAm + CNV + RPPA | 0.84 | 0.82 |
| | miRNA + CNV + RPPA | 0.82 | 0.81 |
| | mRNA + DNAm + CNV | 0.88 | 0.86 |
| | miRNA + DNAm + CNV | 0.82 | 0.81 |
| | mRNA + DNAm + RPPA | 0.88 | 0.86 |
| | mRNA + miRNA + CNV + RPPA | 0.88 | 0.86 |
| | mRNA + miRNA + DNAm + CNV | 0.89 | 0.86 |
| | mRNA + miRNA + DNAm + RPPA | 0.89 | 0.87 |
| | mRNA + DNAm + CNV + RPPA | 0.89 | 0.87 |
| | miRNA + DNAm + CNV + RPPA | 0.85 | 0.83 |
| | mRNA + miRNA + DNAm + CNV + RPPA | 0.89 | 0.88 |

**Table 5.** BRCA Component Performance Fold Summary

| BRCA | | | | | | |
|---|---|---|---|---|---|---|
| | PSN + MME | | PSN | | MME | |
| | Accuracy | F1 | Accuracy | F1 | Accuracy | F1 |
| Fold 1 | 0.903 | 0.884 | 0.871 | 0.839 | 0.857 | 0.833 |
| Fold 2 | 0.894 | 0.870 | 0.88 | 0.85 | 0.829 | 0.795 |
| Fold 3 | 0.871 | 0.857 | 0.899 | 0.885 | 0.834 | 0.816 |
| Fold 4 | 0.889 | 0.869 | 0.907 | 0.886 | 0.843 | 0.82 |
| Fold 5 | 0.907 | 0.890 | 0.861 | 0.83 | 0.843 | 0.821 |

**Table 6.** LGG Component Performance Fold Summary

| LGG | | | | | | |
|---|---|---|---|---|---|---|
| | PSN + MME | | PSN | | MME | |
| | Accuracy | F1 | Accuracy | F1 | Accuracy | F1 |
| Fold 1 | 0.924 | 0.907 | 0.902 | 0.882 | 0.721 | 0.718 |
| Fold 2 | 0.913 | 0.895 | 0.924 | 0.909 | 0.685 | 0.662 |
| Fold 3 | 0.857 | 0.833 | 0.912 | 0.896 | 0.703 | 0.674 |
| Fold 4 | 0.934 | 0.92 | 0.879 | 0.856 | 0.648 | 0.672 |
| Fold 5 | 0.89 | 0.868 | 0.901 | 0.883 | 0.681 | 0.652 |

**Table 7.** KIPAN Component Performance Fold Summary

| KIPAN | | | | | | |
|---|---|---|---|---|---|---|
| | PSN + MME | | PSN | | MME | |
| | Accuracy | F1 | Accuracy | F1 | Accuracy | F1 |
| Fold 1 | 0.955 | 0.945 | 0.966 | 0.956 | 0.972 | 0.962 |
| Fold 2 | 0.961 | 0.949 | 0.966 | 0.956 | 0.972 | 0.964 |
| Fold 3 | 0.961 | 0.951 | 0.978 | 0.971 | 0.961 | 0.954 |
| Fold 4 | 0.96 | 0.951 | 0.955 | 0.943 | 0.994 | 0.993 |
| Fold 5 | 0.955 | 0.942 | 0.966 | 0.955 | 0.977 | 0.971 |

**Table 8.** TCGA DNAm Over Representation Analysis Results

| Dataset | Gene set | Term | Overlap | P-value | Adjusted P-value | Odds Ratio |
|---|---|---|---|---|---|---|
| BRCA | MSigDB Hallmark 2020 | Estrogen Response Early | 100/200 | 1.02445e-10 | 5.12225e-09 | 2.53256 |
| | | Epithelial Mesenchymal Transition | 95/200 | 8.87321e-09 | 2.2183e-07 | 2.28852 |
| | | Myogenesis | 91/200 | 2.20846e-07 | 3.68076e-06 | 2.10961 |
| | KEGG 2021 Human | Axon guidance | 103/182 | 1.7986e-15 | 5.71956e-13 | 3.3086 |
| | | Proteoglycans in cancer | 108/205 | 2.52725e-13 | 4.01832e-11 | 2.82438 |
| | | Pathways in cancer | 227/531 | 9.28599e-13 | 7.96066e-11 | 1.90712 |
| KIPAN | MSigDB Hallmark 2020 | Estrogen Response Early | 106/200 | 1.19312e-11 | 5.96561e-10 | 2.63818 |
| | | Myogenesis | 98/200 | 1.62511e-08 | 4.06277e-07 | 2.24345 |
| | | Epithelial Mesenchymal Transition | 93/200 | 7.80117e-07 | 9.75147e-06 | 2.02706 |
| | KEGG 2021 Human | Axon guidance | 110/182 | 2.22215e-17 | 7.04421e-15 | 3.58235 |
| | | Proteoglycans in cancer | 113/205 | 7.72699e-14 | 1.22473e-11 | 2.87735 |
| | | Human papillomavirus infection | 163/331 | 1.92237e-13 | 2.0313e-11 | 2.2797 |
| LGG | MSigDB Hallmark 2020 | Estrogen Response Early | 108/200 | 2.62026e-11 | 1.31013e-09 | 2.58899 |
| | | Epithelial Mesenchymal Transition | 104/200 | 1.05084e-09 | 2.62711e-08 | 2.38698 |
| | | Apical Junction | 98/200 | 1.47716e-07 | 2.46193e-06 | 2.11397 |
| | KEGG 2021 Human | Axon guidance | 113/182 | 1.28249e-17 | 4.0783e-15 | 3.62084 |
| | | Focal adhesion | 121/201 | 3.05032e-17 | 4.85001e-15 | 3.34571 |
| | | Proteoglycans in cancer | 122/205 | 8.00606e-17 | 8.48642e-15 | 3.25124 |

**Table 9.** TCGA mRNA Gene Set Enrichment Analysis Results

| Dataset | Gene set | Term | ES | NES | NOM p-val | FDR q-val | Tag % | Gene % |
|---|---|---|---|---|---|---|---|---|
| BRCA | KEGG 2021 Human | Pentose and glucuronate interconversions | -0.774355 | -2.29015 | 0 | 0 | 10/16 | 9.58% |
| | | Cell cycle | 0.573984 | 2.27577 | 0 | 0 | 48/108 | 16.26% |
| | | Homologous recombination | -0.649829 | -2.39384 | 0 | 0 | 13/25 | 14.04% |
| | MSigDB Hallmark 2020 | G2-M Checkpoint | 0.576235 | 2.46117 | 0 | 0 | 102/188 | 23.18% |
| | | Androgen Response | -0.499048 | -2.03848 | 0 | 0.00577345 | 37/68 | 25.03% |
| | | Myc Targets V1 | 0.446703 | 1.88204 | 0 | 0.00801285 | 129/175 | 46.57% |
| LGG | KEGG 2021 Human | Nicotine addiction | 0.80602 | 3.10307 | 0 | 0 | 15/18 | 11.28% |
| | | Systemic lupus erythematosus | -0.641083 | -2.29704 | 0 | 0 | 35/52 | 20.32% |
| | | Type I diabetes mellitus | -0.737799 | -2.2463 | 0 | 0 | 19/24 | 15.40% |
| | MSigDB Hallmark 2020 | G2-M Checkpoint | -0.672679 | -2.6506 | 0 | 0 | 53/126 | 8.53% |
| | | E2F Targets | -0.641025 | -2.52297 | 0 | 0 | 66/137 | 15.34% |
| | | Epithelial Mesenchymal Transition | -0.621457 | -2.45649 | 0 | 0 | 94/144 | 23.64% |
| KIPAN | KEGG 2021 Human | Oxidative phosphorylation | 0.623763 | 3.14138 | 0 | 0 | 87/112 | 24.92% |
| | | Pentose and glucuronate interconversions | -0.668482 | -2.11695 | 0 | 0 | 11/24 | 10.08% |
| | | Intestinal immune network for IgA production | -0.624027 | -2.15917 | 0 | 0 | 31/40 | 27.41% |
| | MSigDB Hallmark 2020 | Oxidative Phosphorylation | 0.604961 | 3.25895 | 0 | 0 | 145/196 | 27.66% |
| | | Allograft Rejection | -0.562653 | -2.43625 | 0 | 0 | 118/178 | 27.41% |
| | | Epithelial Mesenchymal Transition | -0.527188 | -2.26799 | 0 | 0 | 111/166 | 30.88% |
